# Impacts of a relational leadership development program on psychological sense of community

**DOI:** 10.1101/2025.02.21.25322652

**Authors:** Naseeha Islam, Senta G. Wiederholt, Brian Park, Anaïs Tuepker, Elaine Waller Uchison, Leah Gordon, Matthew Lewis, Alexander Mansour, Nicole A. Steckler, Samuel T. Edwards

## Abstract

**Introduction:** In a time of increasing employee dissatisfaction and burnout within healthcare, relational leadership approaches have the potential to improve belonging and a sense of community among the workforce.

**Methods:** The Relational Leadership Institute (RLI) teaches a relationship-based leadership model to help participants enhance interpersonal skills in the healthcare workplace. A survey was conducted of participants to assess their psychological sense of community (PSOC) throughout this leadership program, at baseline, post-program, and 6-months post-program.

**Results:** Overall mean PSOC was 4.87 (SD 0.68) at baseline, 5.16 (SD 0.77) post-program, and 5.13 (SD 0.72) 6-months post-program, with a significant increase from baseline to post-program (p<0.01) and baseline to 6-months post-program (p=0.01). There was no significant difference between post-program and 6-months post-program PSOC levels (p=0.99). Baseline mean PSOC score was greater among non-white participants (mean 5.00 vs. 4.83 among white participants, p<0.01), and greater among trainees than among working professionals (mean 4.96 vs. 4.82, p<0.01).

**Conclusion:** The RLI intervention has a significant impact on the psychological sense of community among participants, suggesting that similar relationship-based programs could support today’s healthcare workforce.

## Introduction

The desire to belong is a strong driver of human motivation.^1^ In the workplace, a sense of belonging is associated with higher employee engagement and lower burnout rates.^2,3^ Relational leadership^4^ – an approach to leadership that brings intention to enhancing team and organizational relationships –-has the potential to build a sense of belonging and community. Relational leadership development programs emphasize the importance of shared values and collective action^5^ and have demonstrated positive impacts on skill retention, individual wellbeing, and psychological safety in the context of interdependent work.^6^ In this study, we investigated how participation in a relational leadership development program impacted psychological sense of community (PSOC), a validated measure assessing an individual’s sense of belonging in a group.^7^

## Methods

The Relational Leadership Institute (RLI) was launched in 2017 at Oregon Health and Science University (OHSU) in Portland, OR. RLI engages participants across a diverse array of healthcare fields to build skills in a relationship-based leadership model.^8^ Over its 3-month, cohort-based curriculum, participants learn evidence-based skills to cultivate psychological safety, narrow power differences, and promote authenticity. RLI is conducted twice per year for approximately 30 participants, and the 10-module curriculum includes sessions on psychological safety, positionality, teaming across differences, strengths-based coaching, and conflict transformation. The curriculum combines large group didactics, small group practice, and skill application in participants’ work environments.

To assess whether RLI impacts participant’s PSOC, we conducted surveys of RLI participants from 2018-2021. Surveys were solicited from participants at baseline (after the introductory session), immediately after the last session of the program, and 6-months after program completion. To assess member’s PSOC, defined as a “supportive network or social structure that one can depend on for psychological significance and identification” we used a validated scale developed by Jason et al.^9^ This 9-item measure included questions categorized into three theoretical subdomains: entity (a group’s organization and purpose), membership (social relationships), and self (identity and importance of self).^9^ We report overall PSOC and subdomains as linear scales with ranges of 1-6, with higher scores indicating greater sense of psychological safety. Participants also self-reported their age, race, training stage, gender, and profession in these surveys, which were analyzed using descriptive statistics and explored for possible interactions and effects on PSOC score using ANOVA. All demographic variables were also explored for potential inclusion in multivariate linear regression models comparing participant PSOC score at each survey time points. All analyses were conducted using RStudio, v.2024.9.0.375.^9^

## Results

Of 192 participants, survey response rates were 86.5% (n=166) at baseline, 67.7% (n=130) post-program, and 44.8% (n=86) 6-months post-program. Table 1 shows a summary of participant demographic data at baseline. Participants reported a mean age of 37.9 years (SD 9.9 years), and predominantly identified as female (84.9%) and white (72.3%). 33.7% of participants were trainees (e.g., residents, students), while 65.1% were working professionals (e.g., nurses, physicians). Baseline mean PSOC score overall was 4.87, greater among non-white participants (mean 5.00 vs. 4.83 among white participants, p<0.01), and greater among trainees than among working professionals (mean 4.96 vs. 4.82, p<0.01). Our analyses found no difference in the change in PSOC scores by age, gender, race, training stage, or profession.

**Table 1.**
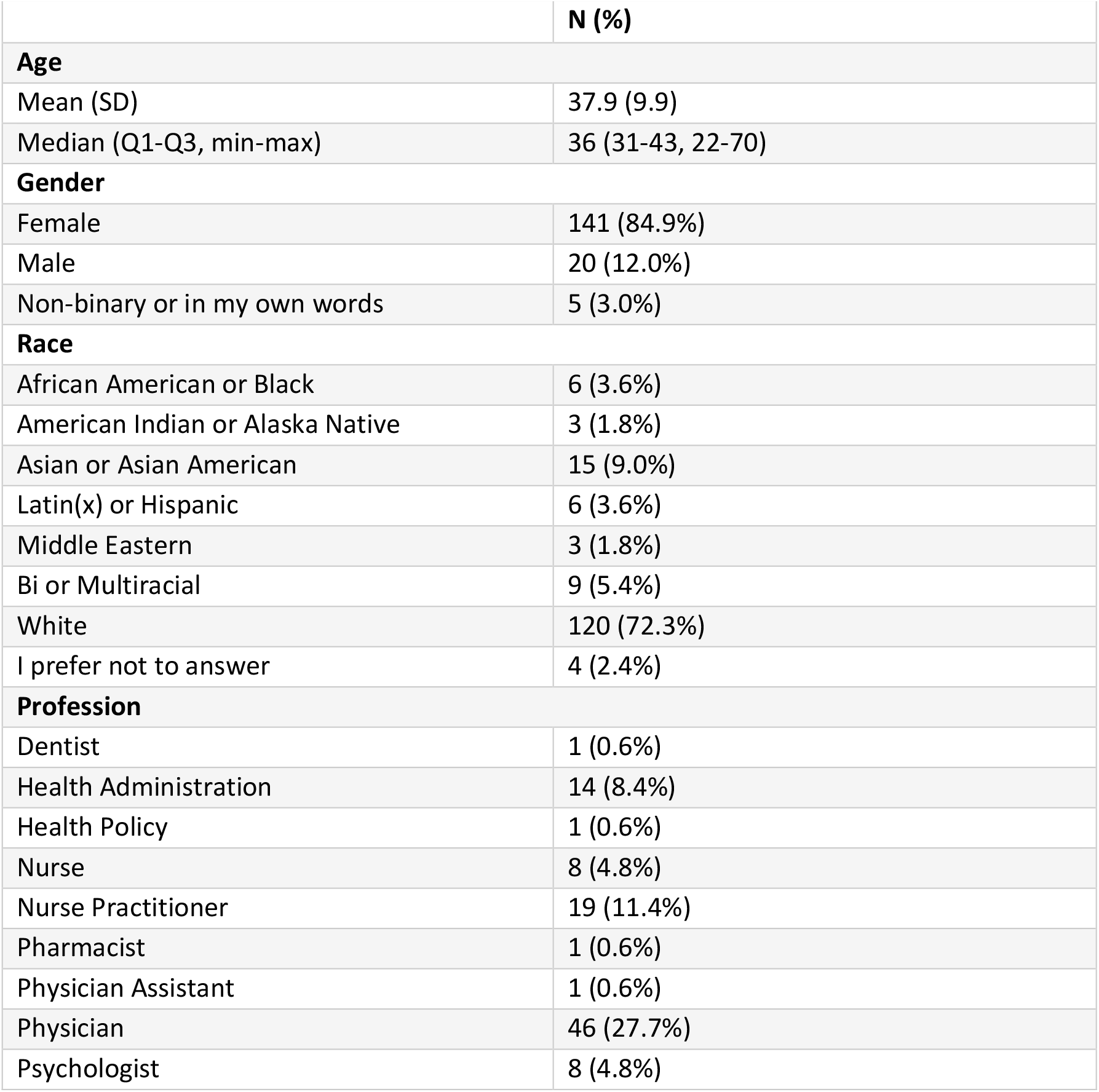
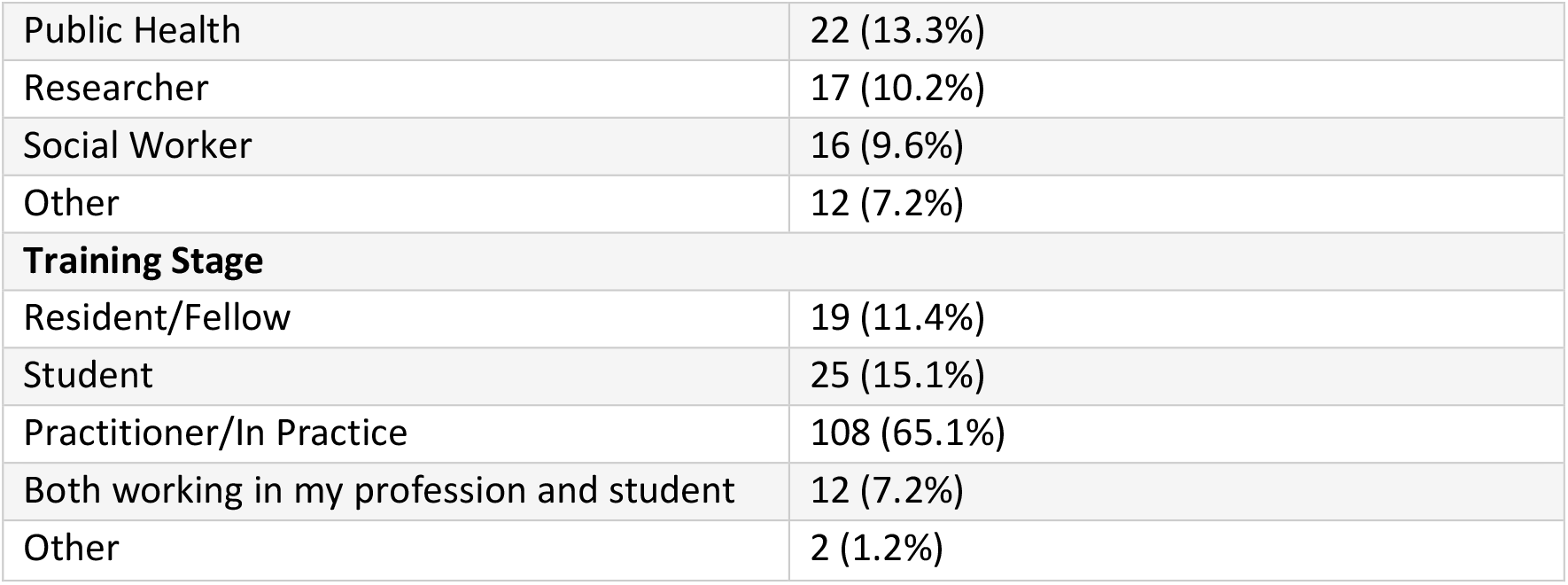
RLI Participant Characteristics at Baseline (N = 166)

|  | N (%) |
| --- | --- |
| **Age** |  |
| Mean (SD) | 37.9 (9.9) |
| Median (Q1-Q3, min-max) | 36 (31-43, 22-70) |
| **Gender** |  |
| Female | 141 (84.9%) |
| Male | 20 (12.0%) |
| Non-binary or in my own words | 5 (3.0%) |
| **Race** |  |
| African American or Black | 6 (3.6%) |
| American Indian or Alaska Native | 3 (1.8%) |
| Asian or Asian American | 15 (9.0%) |
| Latin(x) or Hispanic | 6 (3.6%) |
| Middle Eastern | 3 (1.8%) |
| Bi or Multiracial | 9 (5.4%) |
| White | 120 (72.3%) |
| I prefer not to answer | 4 (2.4%) |
| **Profession** |  |
| Dentist | 1 (0.6%) |
| Health Administration | 14 (8.4%) |
| Health Policy | 1 (0.6%) |
| Nurse | 8 (4.8%) |
| Nurse Practitioner | 19 (11.4%) |
| Pharmacist | 1 (0.6%) |
| Physician Assistant | 1 (0.6%) |
| Physician | 46 (27.7%) |
| Psychologist | 8 (4.8%) |
| Public Health | 22 (13.3%) |
| Researcher | 17 (10.2%) |
| Social Worker | 16 (9.6%) |
| Other | 12 (7.2%) |
| **Training Stage** |  |
| Resident/Fellow | 19 (11.4%) |
| Student | 25 (15.1%) |
| Practitioner/In Practice | 108 (65.1%) |
| Both working in my profession and student | 12 (7.2%) |
| Other | 2 (1.2%) |

Figure 1 shows change in adjusted means in overall PSOC and PSOC subdomains. Overall mean PSOC was 4.87 (SD 0.68) at baseline, 5.16 (SD 0.77) post-program, and 5.13 (SD 0.72) 6-months post-program. In regression analyses adjusted for race (white vs. non-white) and trainee stage (trainee vs. professional), mean PSOC was significantly different from baseline to post-program at p<0.01, and this improvement was sustained at six months (baseline vs. 6-month post-program, p=0.01). There was no significant difference between post-program and 6-month post-program PSOC levels (p=0.99). Stratified analyses by PSOC demonstrated similar increases in the member and self subdomains, but no change in the entity domain.

**Figure 1.**
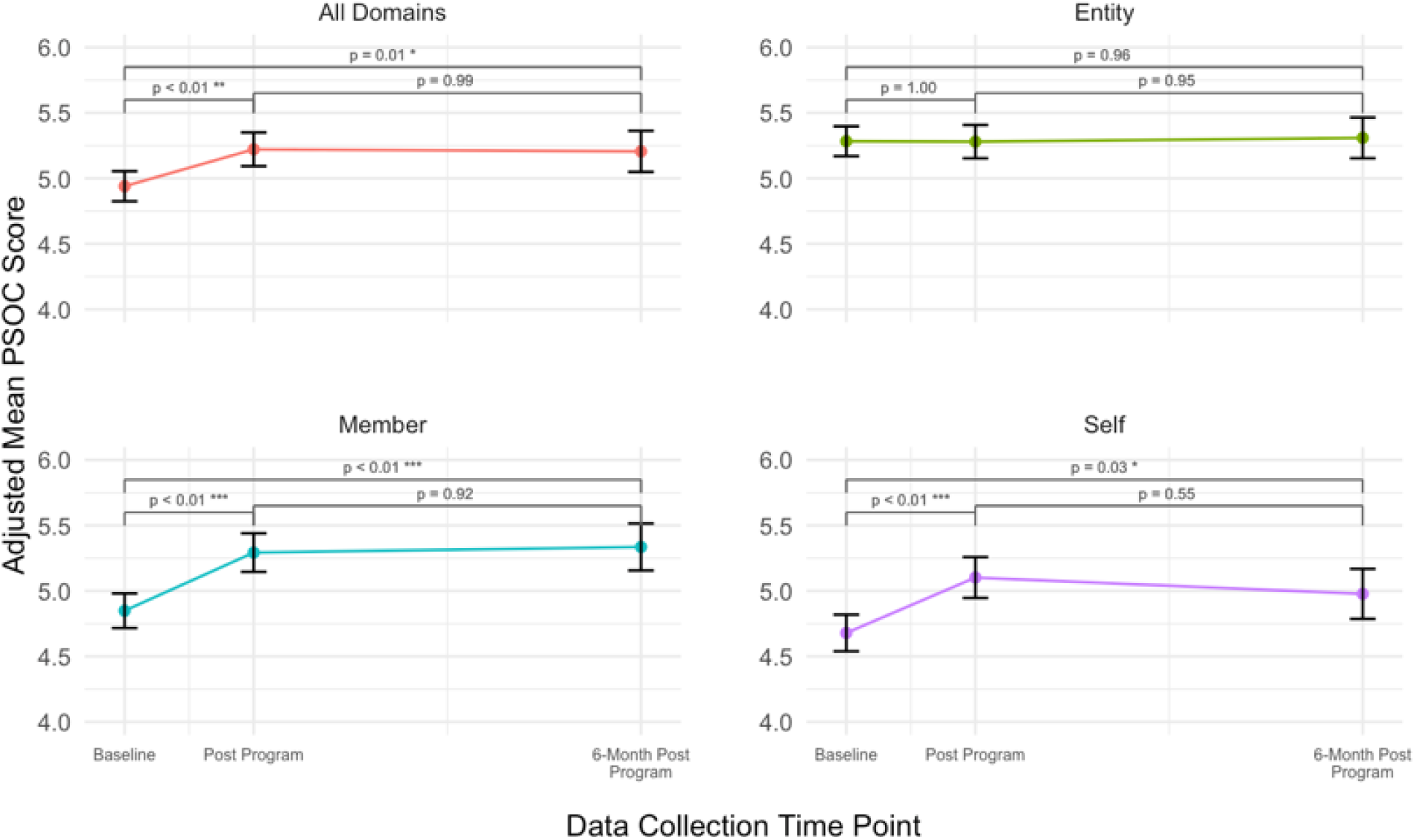
Change in Adjusted Means in Overall PSOC and PSOC Subdomains

## Discussion

This study assessed the impacts a relational leadership program had on PSOC across 3 years of participants. We found that the program significantly improves participants’ PSOC from pre- to post-program, with these enhancements sustaining out to 6-months post-program. In conjunction with prior studies that demonstrated that interventions designed to enhance relational leadership capacity may enhance application of relational leadership skills and participant well-being, this study suggests programs like RLI may also support healthcare professionals and trainees to build and develop a protective sense of belonging. Given prior research that has demonstrated associations between enhanced PSOC and a sense of well-being, these findings have significant implications for healthcare systems, during a time when dissatisfaction, burnout, and employee turnover are at record highs. Similar programs focused on relational leadership development could be implemented at other institutions to foster interprofessional community, to curb burnout and enhance a sense of belonging in the workplace.

The findings that participants of color exhibited significantly higher baseline PSOC scores than their white counterparts, and that the intervention appeared to improve PSOC scores equally between demographics, are notable. Previous studies have shown that individuals whose racial identities are under-represented in medicine (i.e., Black, Indigenous, Latinx, Pacific Islander) experience significantly higher rates of burnout^3^ and isolation^10^ than their counterparts who are not under-represented. Our study suggests that relational leadership programs hold equal potential to enhance a sense of community among groups within healthcare that often disproportionately experience isolation and burnout. More research is necessary, however, to better understand the mechanisms by which relational leadership programs may foster community to the benefit of under-represented individuals. Additionally, baseline PSOC scores were significantly higher for participants in training (versus their professional counterparts). Prior research has suggested that healthcare training can often be isolating and/or morally injurious; our results demonstrate an equal increase in PSOC score between trainees and professionals after the intervention, and sustainment at 6 months. These findings suggest that relational leadership development programs may provide protection for healthcare trainees from future burnout in the field, a hypothesis that could be explored further in future research.^10^

One limitation of this study is the decreasing response rate of participants at progressing time points. Incomplete participant responses, especially in the context of building community in professional settings, obscure the full impacts of a relational leadership intervention on participants’ PSOC. Additionally, skewed demographic proportions of participants (e.g., more white participants than those holding minoritized identities) may confound results.

## Conclusions

RLI is a leadership development program that emphasizes relational skills across a range of health professionals at varying stages. We found there to be a significant impact on the psychological sense of community among participants. Findings were especially encouraging of the enduring impacts of such programs, and the potential to improve experiences of groups that may be disproportionately harmed in the healthcare workforce (i.e., trainees, underrepresented identities). Overall, our study provides support for similar relationship-based interventions to develop and shift the culture of our current healthcare industry.

## Data Availability

All data produced in the present study are available upon reasonable request to the authors

## Ethics

Ethical approval for this study regarding human subjects research was provided by the Institutional Review Board at Oregon Health & Science University.

